# Patterns of substance use during pregnancy are differentially associated with maternal mental health across the perinatal period

**DOI:** 10.64898/2026.10.01.26364530

**Authors:** Kimberly N. Palmer, Ka Ying Toby Law, Suena H. Massey, Alexandre A. Lussier

## Abstract

**Background:** Substance use during pregnancy can substantially increase risk for postpartum depression and anxiety symptoms, which can have lasting effects on family functioning and child development. However, substance use is often conceptualized as a binary exposure, limiting our understanding of whether specific patterns of use are associated with maternal mental health in the perinatal period. Here, we examined longitudinal patterns of prenatal alcohol use and smoking in relation to maternal depressive and anxiety symptoms across pregnancy and the first two years postpartum.

**Methods:** We analyzed prospective data from the Avon Longitudinal Study of Parents and Children (ALSPAC), a population-based cohort from the UK. Alcohol and cigarette use were assessed across all three trimesters and modeled as longitudinal patterns (abstained, persistent, intermittent, discontinued). Maternal depressive and anxiety symptoms were measured from the second trimester through 21 months postpartum using the Crown-Crisp Experiential Index.

**Results:** Persistent alcohol use was associated with higher depressive and anxiety symptoms and slower symptom declines over time, while women with intermittent or discontinued use had symptom profiles similar to abstainers. By contrast, persistent, intermittent, and quitting smoking patterns were associated with higher symptom levels overall, with persistent smoking additionally associated with steeper declines in symptoms over time.

**Conclusion:** Our findings highlight the importance of considering the persistence and cessation of substance use during pregnancy in the context of maternal mental health, pointing to patterns of persistent substance use as a potential target to inform screening and early intervention for women at higher risk for perinatal psychological distress.

## INTRODUCTION

Maternal mental health during pregnancy and the postpartum period plays a central role in shaping how women navigate the physiological, emotional, and social transitions of early motherhood. Depression and anxiety are among the most common complications of pregnancy, affecting up to one in five women, and are associated with impaired daily functioning, disrupted sleep, and reduced overall well-being (Gavin et al., 2005; Leach et al., 2016; Woody et al., 2017). These symptoms do not emerge in a vacuum but unfold within a period marked by major hormonal changes, shifting identities, and evolving social roles, many of which continue well beyond delivery (Satyanarayana et al., 2011). As a result, emotional distress that begins during pregnancy often persists into the postpartum period rather than resolving spontaneously. Postpartum depression, in particular, is a substantial public health concern, with prevalence estimates consistently ranging between 10% and 20% worldwide (Liu et al., 2021). Importantly, these postpartum symptoms frequently reflect vulnerabilities that are already present during pregnancy.

Recognition of this continuity has shifted attention toward the prenatal period as a key window for identifying risk factors for postpartum depression and anxiety. Evidence from large meta-analyses indicates depression during pregnancy and prior mental health history are among the strongest predictors of postpartum depressive symptoms, often outweighing the influence of obstetric or demographic factors (Liu et al., 2021; Qiu et al., 2022). These findings complicate narratives that frame postpartum depression as a sudden or unexpected response to childbirth.

Instead, these findings suggest that risk for postpartum psychopathology often develops earlier and follows a continuous trajectory across the perinatal transition. Viewed in this way, pregnancy is a key window to observe early behavioral and clinical indicators that may signal heightened vulnerability for persistent psychological distress after birth.

One behavioral indicator that has received increasing attention in this context is substance use during pregnancy. Across multiple meta-analyses, prenatal substance use has been associated with more than threefold higher odds of postpartum depression, although the strength of this association varies by substance type (Pacho et al., 2023; Qiu et al., 2022). Within this literature, the prevalence of postpartum depression varies by substance type, with estimates of approximately 23% among women reporting alcohol use, 27% among those using tobacco, and up to 44% among those using multiple substances during pregnancy (Pacho et al., 2023). Overall, these findings suggest prenatal substance use may reflect broader vulnerability to postpartum psychopathology, rather than functioning as a discrete health behavior independent of underlying psychosocial and mental health risk.

Despite this accumulating evidence, prior studies most often conceptualize prenatal substance use as a binary exposure, classifying women as users or non-users (Estabrook et al., 2016). Such classifications obscure meaningful variation in how substance use changes across pregnancy and into the postpartum period, overlooking the fact that some women discontinue use following pregnancy recognition, others reduce use inconsistently, and some persist throughout pregnancy (Forray, 2016; Massey et al., 2021). These distinct patterns likely reflect heterogeneity in vulnerability, available resources, and life circumstances, pointing to substance use as a dynamic process, as opposed to a static behavior. Treating substance use as fixed exposures therefore limits our ability to distinguish women whose risk profiles evolve over time from those whose risk remains stable. Longitudinal assessment of substance use patterns offers a way to address this limitation by capturing within-person change and aligning analytic approaches with the temporal complexity of pregnancy (Massey et al., 2021).

Importantly, prenatal substance exposure classifications also encode maternal behavior. Women who discontinue, reduce, intermittently use, or persist in substance use during pregnancy may differ in ways that are obscured when exposure is represented as a binary variable. Prior work has demonstrated that more granular characterization of prenatal substance use can alter estimates of both maternal correlates and offspring risk, underscoring the importance of distinguishing patterns of maternal behavior rather than exposure status alone (Massey et al., 2021). However, research examining these patterns alongside concurrent trajectories of depressive and anxiety symptoms remains limited, leaving it unclear whether changes in substance use across pregnancy could help identify women experiencing psychological distress that persists into the postpartum period.

The present study seeks to address this gap by examining how patterns of alcohol and cigarette use across pregnancy relate to maternal depressive and anxiety symptoms throughout the postpartum period. Rather than treating substance use as a binary prenatal exposure, we modeled longitudinal patterns of consumption across pregnancy (abstainers, quitters, intermittent users, and persistent users), representing observable indicators of vulnerability that may help identify women at elevated risk for sustained psychological distress. Specifically, we hypothesize that intermittent and persistent substance use is associated with higher and more persistent levels of depressive and anxiety symptoms from trimester two of pregnancy through 21 months postpartum. By contrast, we expect discontinuation during pregnancy to align with more favorable mental health trajectories, similar to those observed among abstainers. By modeling substance use patterns and mental health symptoms jointly, this study aims to clarify how prenatal behaviors signal risk for postpartum mental health outcomes and to inform earlier identification of women who may benefit from targeted perinatal support.

## METHODS

### Study design and sample

We analyzed prospective data from the Avon Longitudinal Study of Parents and Children (ALSPAC), a population-based birth cohort that recruited pregnant women residing in Avon, United Kingdom, with expected delivery dates between 1 April 1991 and 31 December 1992 (Boyd et al., 2013; Fraser et al., 2013; Major-Smith et al., 2022). A total of 20,248 pregnancies were identified as eligible, of which 14,541 were enrolled, resulting in 14,062 live births and 13,988 children alive at 1 year. The study website provides details of all collected and generated data, which are available through a fully searchable data dictionary and variable search tool https://www.bristol.ac.uk/alspac/researchers/our-data/. The present study focused on mothers with available data on prenatal substance use and mental health outcomes.

### Prenatal substance use patterns

Alcohol use and cigarette smoking were assessed via self-administered questionnaires completed at prenatal timepoints corresponding to the first, second, and third trimesters of pregnancy. Alcohol consumption and smoking behavior were reported at each trimester and respectively recoded into binary indicators of any use (1) versus no use (0). See details on coding of trimester-specific timepoints in **Supplemental Materials**.

Using these trimester-specific indicators, longitudinal patterns of prenatal substance use were constructed separately for alcohol and smoking. Based on repeated assessments, four mutually exclusive patterns were defined to capture variation in the timing and consistency of prenatal substance use. Participants with missing data from two or more trimesters were excluded from our analyses. These pattern variables were used as the primary exposure measures in subsequent analyses examining associations with depressive and anxiety symptoms from pregnancy through 21 months postpartum (**Figure S1**).

● ***Abstainers:*** No reported substance use (0) at any trimester-specific timepoint.
● ***Persistent users:*** Reported substance use (1) at all trimester-specific timepoints.
● ***Intermittent users:*** Reported substance use (1) at one or more trimester-specific timepoints, but not consistently across all trimesters.
● ***Quitters:*** Reported substance use (1) during the first trimester, followed by no use (0) at subsequent trimester-specific timepoints.

### Maternal psychopathology

Maternal depressive and anxiety symptoms were assessed during the second and third trimesters of pregnancy and at 8 weeks, 8 months, and 21 months postpartum using standardized self-report instruments. Anxiety and depressive symptoms were assessed using subscales from the Crown–Crisp Experiential Index (CCEI), which captures core dimensions of affective distress, including generalized anxiety, somatic anxiety, and depressive symptoms (Crown & Crisp, 1979). Each subscale consists of eight dichotomously scored items, yielding total scores ranging from 0 to 8, with higher scores indicating higher levels of symptoms. The CCEI has been widely used in perinatal populations and demonstrates strong psychometric properties, including good internal consistency and construct validity, within the ALSPAC cohort (Evans et al., 2001).

### Covariates

Maternal and demographic characteristics associated with prenatal substance use and maternal mental health were included as covariates in all analyses. Covariates included maternal age at delivery, maternal education, marital status, parity, maternal race/ethnicity, and infant sex. See details on selection and coding of covariates in **Supplemental Materials**.

### Statistical analyses

Our primary analytic sample included 7,984 mothers with complete data on both maternal substance use patterns and covariates, with a child alive at 1 year. Mothers of twins were also excluded (**Table 1**). Prenatal alcohol and smoking pattern variables were treated as categorical predictors, with abstainers/non-users specified as the reference group in all analyses.

**Table 1:** Sample characteristics of the analytic sample and full ALSPAC cohort.

| **Variable** | **ALSPAC sample**<br>**N=13,646 (%)** | **Analytic sample**<br>**N=7,984 (%)** |
| --- | --- | --- |
| **Age** |  |  |
| 15-19 years | 648 (4.7%) | 230 (2.9%) |
| 20-35 years | 12,000 (87.9%) | 7,163 (89.7%) |
| 36+ years | 942 (6.9%) | 591 (7.4%) |
| missing | 56 (0.4%) | - |
| **Education** |  |  |
| Less than O-level | 3,632 (26.6%) | 2,106 (26.4%) |
| O-level | 4,174 (30.6%) | 2,890 (36.2%) |
| A-level | 2,720 (23.8%) | 1,902 (23.8%) |
| Degree or above | 1,550 (11.4%) | 1,086 (13.6%) |
| missing | 1,570 (11.5%) | - |
| **Race/Ethnicity** |  |  |
| White | 11,677 (85.6%) | 7,825 (98%) |
| Non-White | 312 (2.3%) | 159 (2%) |
| missing | 1,657 (12.1%) | - |
| **Previous pregnancies** |  |  |
| 0 | 5,649 (41.4%) | 3,631 (45.5%) |
| 1 | 4,385 (32.1%) | 2,832 (35.5%) |
| 2 | 1,794 (13.1%) | 1,111 (13.9%) |
| 3+ | 747 (5.9%) | 410 (5.1%) |
| missing | 1,071 (8.5%) | - |
| **Marital status** |  |  |
| Married | 9,539 (46.4%) | 6,268 (78.5%) |
| Never Married | 2,429 (11.8%) | 1,292 (16.2%) |
| Widowed/Divorced/Sep<br>arated | 766 (37.3%) | 424 (5.3%) |
| missing | 912 (4.4%) | - |
| **Sex of the baby** |  |  |
| Male | 7,009 (51.4%) | 4,098 (51.3%) |
| Female | 6,611 (48.4%) | 3,886 (48.7%) |
| missing | 26 (0.2%) | - |

**Note.** Values are presented as counts (n) and percentages (%), and participants who did not
report were categorized as missing. The full ALSPAC cohort refers to mothers of singleton
births, who are in core ALSPAC sample, and whose child is alive at 1 year.

As our first set of analyses, we fit models separately for each outcome (anxiety and depressive symptoms) at each timepoint, adjusting for all covariates. Models were estimated in *R* (version 4.3) using the linear regression function, and results are presented as standardized beta coefficients with 95% confidence intervals. As missingness patterns were different across the outcome timepoints, the specific N for each analysis varied slightly from our primary analytic sample (see **Table 2**).

**Table 2.**
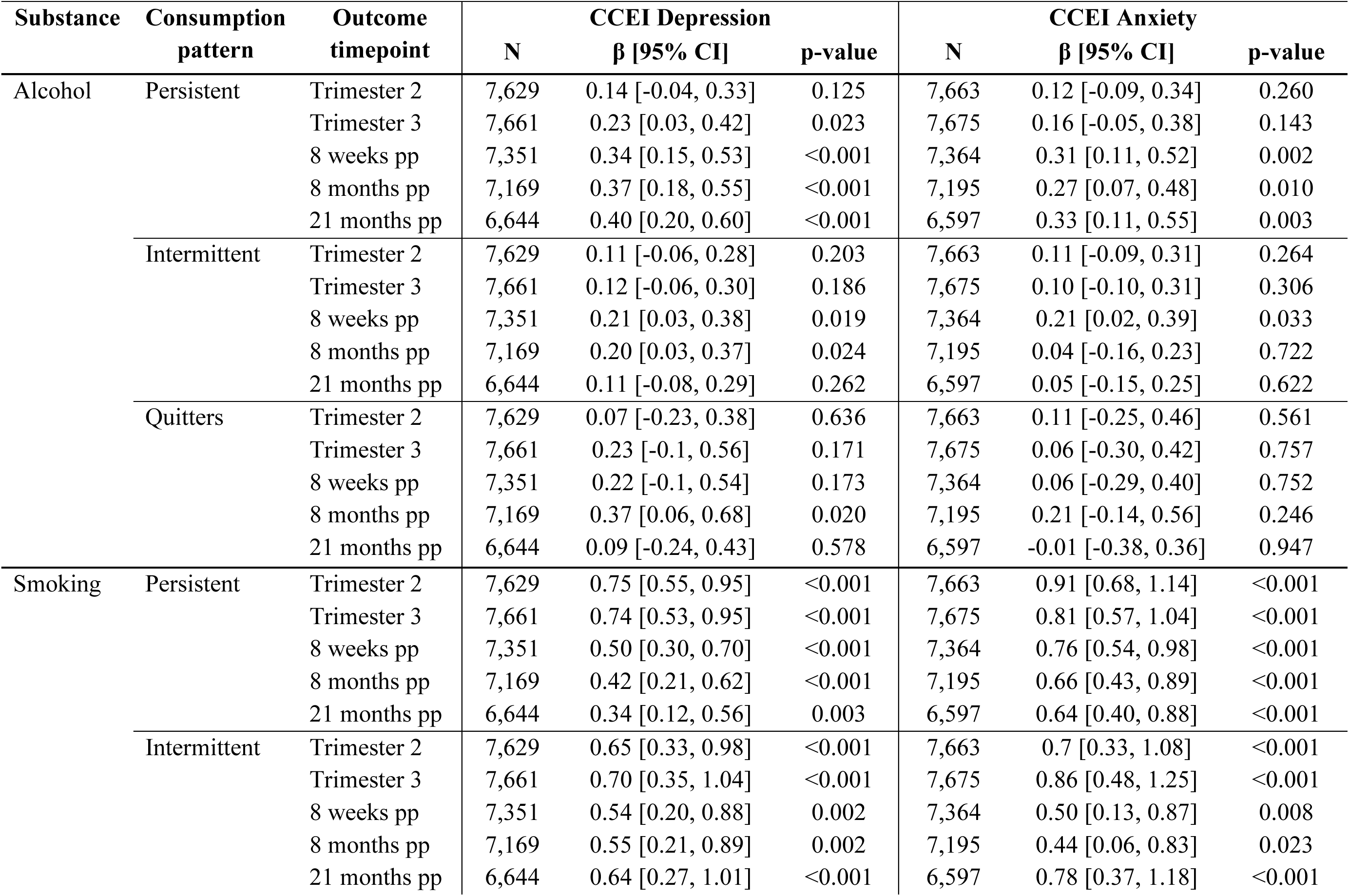
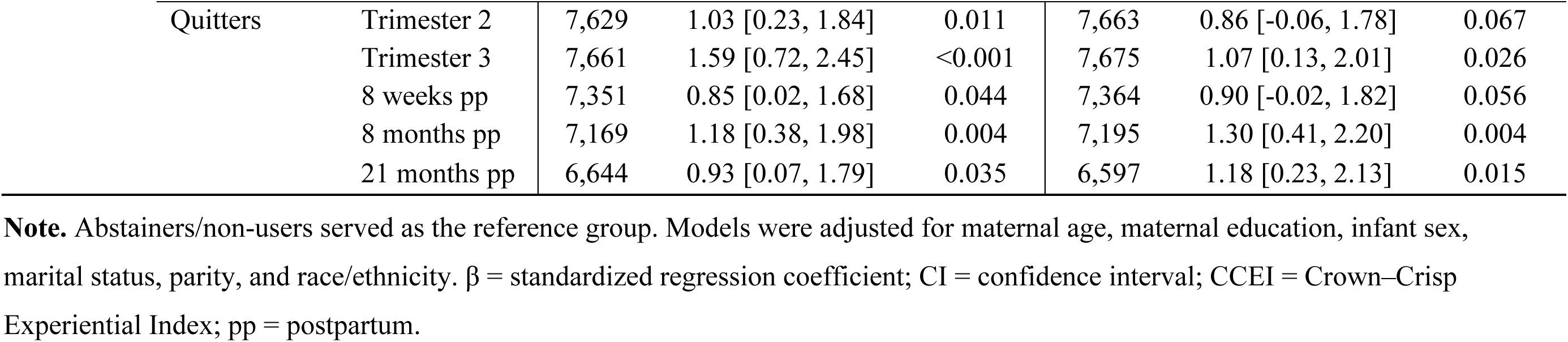
Associations between prenatal consumption patterns and maternal depressive and anxiety symptoms.

Next, we conducted linear mixed-effects modelling to determine whether substance use patterns predicted different trajectories of depressive or anxiety symptoms from trimester 2 to 21 months. We used the *lmer* function of the *lme4* package, adjusting for all covariates and repeated measurements across participants. As linear mixed-effects models account for missingness, we used the entire analytic sample for these analyses (N=7,984).

Finally, we conducted two primary sensitivity analyses. First, we analyzed continued depressive symptom scores measured from the Edinburgh Postnatal Depression Scale (EPDS; see details in Supplemental). Second, we corrected for other substance use patterns (i.e., correcting for smoking patterns in the alcohol analyses, and vice versa).

## RESULTS

### Sample characteristics

**Table 1** compares demographic characteristics of the analytic sample (N=7,984) with those of the full ALSPAC mother cohort (N=13,646). Aside from having a higher proportion of mothers who are currently married, the demographic composition of the analytic sample closely resembled that of the broader ALSPAC cohort, suggesting that the analytic sample remained broadly representative of the larger cohort.

Alcohol use patterns varied across pregnancy, with most participants classified as Abstainers (57.1%). Persistent alcohol use was reported by 17.3% of the sample, and 20.5% were classified as Intermittent users throughout pregnancy. A small proportion reported alcohol use early in pregnancy followed by cessation (Quitters, 5.1%; **Figure 1**).

**Figure 1.**
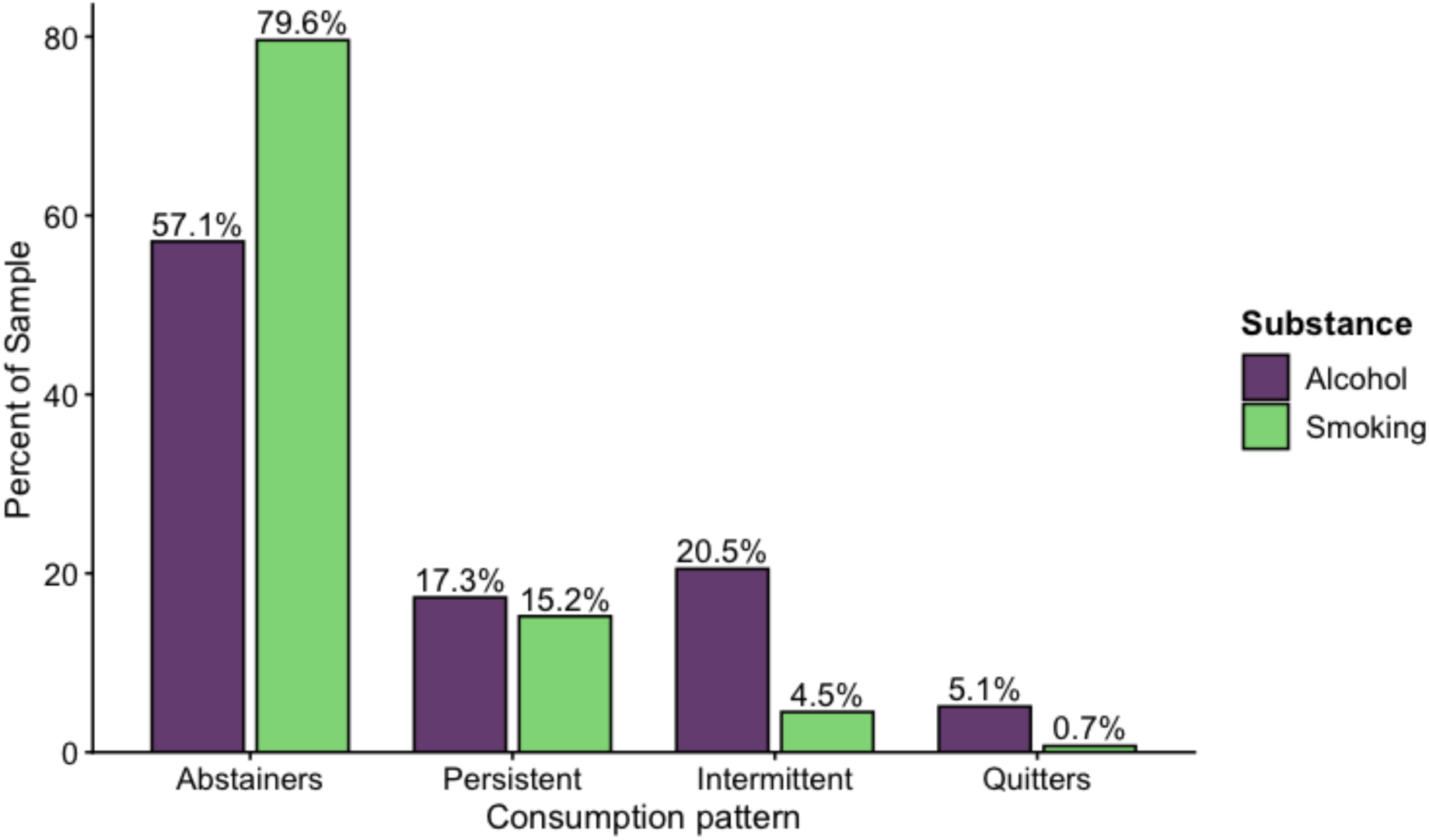
Distribution of prenatal alcohol and smoking use patterns across pregnancy. Percentage of the sample classified as abstainers, persistent, intermittent, and quitters for prenatal alcohol and smoking use patterns across pregnancy. Abstainer patterns indicate no substance use across pregnancy. Persistent patterns indicate continued use across pregnancy, intermittent patterns indicate inconsistent use across pregnancy, and quitter patterns indicate use early in pregnancy followed by discontinuation. Percentages are presented separately for alcohol and smoking use.

Smoking showed a different distribution. The majority of participants were classified as Abstainers (79.6%). Persistent smoking occurred in 15.2% of participants, while 4.5% were classified as intermittent smokers across pregnancy. Quitters only represented 0.7% of the sample (**Figure 1**).

### Alcohol Use Patterns and Psychopathology

Persistent alcohol use was associated with higher CCEI depressive symptoms beginning in the third trimester and extending across the postpartum period (β range=0.23–0.40; p<0.05). Persistent alcohol use was also associated with higher CCEI anxiety symptoms across the postpartum period (β range=0.31–0.33; p<0.05). By contrast, intermittent alcohol use showed modest and inconsistent associations with depressive and anxiety symptoms, with significant associations emerging primarily during the early postpartum period (depression: 8 weeks postpartum, β=0.21, p=0.019; 8 months postpartum, β=0.20, p=0.024; anxiety: 8 weeks postpartum, β=0.21, p=0.033). Participants who quit alcohol use during pregnancy generally did not differ from abstainers across depressive or anxiety outcomes, although quitters reported higher depressive symptoms at 8 months postpartum (β=0.37, p=0.020) (**Table 2**; **Figure 2**).

**Figure 2.**
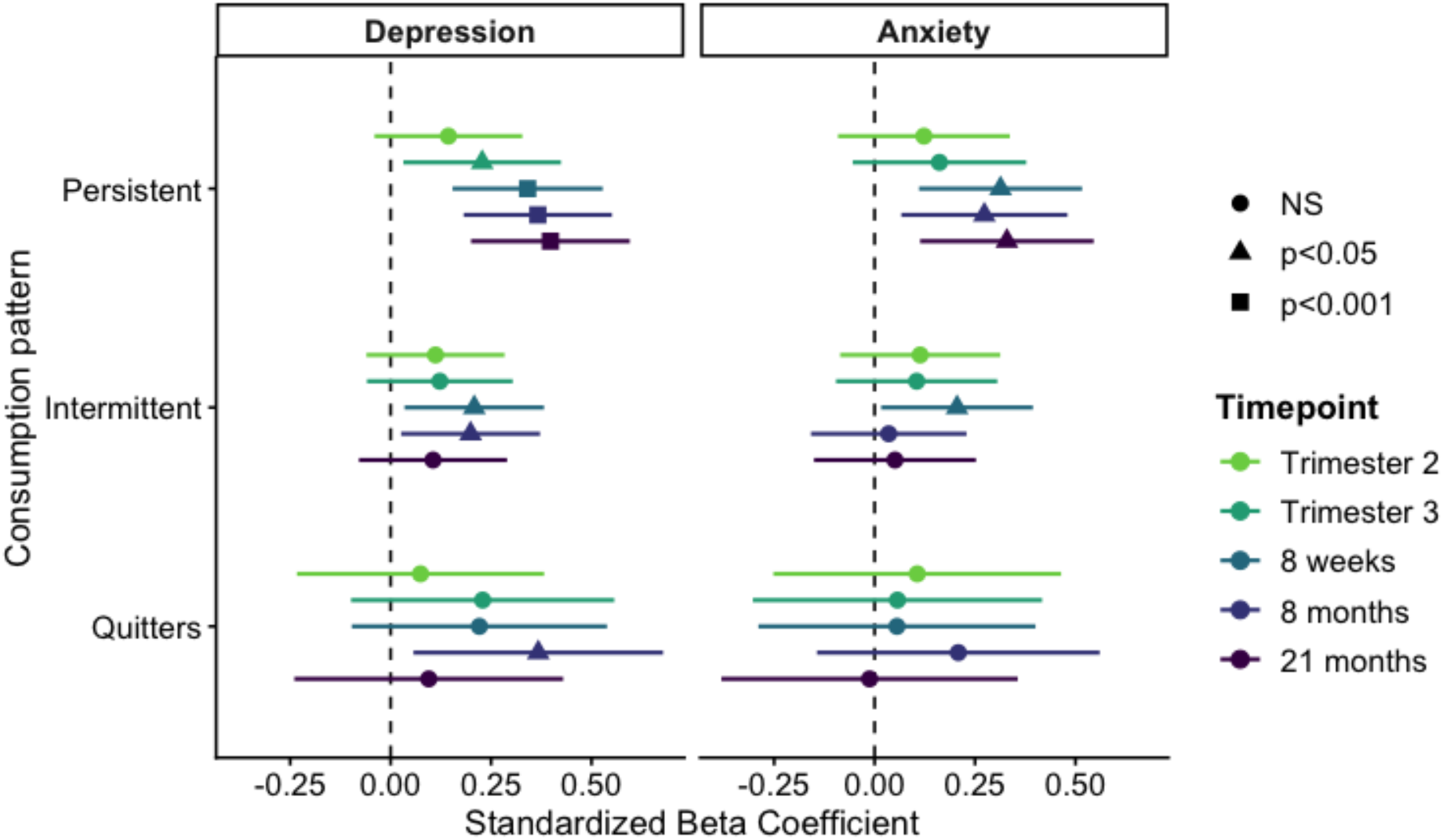
Associations Between Prenatal Alcohol Use Patterns and Maternal Depressive and Anxiety Symptoms Across Pregnancy and Postpartum. Points represent standardized regression coefficients (β), and error bars represent 95% confidence intervals. Abstainers/non-users served as the reference group. Positive coefficients indicate higher symptom levels relative to abstainers. Depressive and anxiety symptoms were measured with the Crown–Crisp Experiential Index (CCEI). 8 weeks, 8 months, and 21 months timepoints were measured after birth.

Longitudinal mixed-effects models revealed a significant effect of time on depressive (β=-0.01, p<0.001) and anxiety symptom levels (β=-0.011, p<0.001), but no main effects of alcohol consumption patterns (**Table 3)**. However, we identified a significant interaction between persistent use and time on both depressive (β=0.003, p<0.001) and anxiety symptoms (β=0.003, p=0.001). In other words, participants with persistent alcohol use across pregnancy showed a slower decline in symptoms across time than abstainers. No interaction was detected for intermittent use or quitters.

**Table 3.** Linear mixed-effect models of consumption patterns and depressive or anxiety symptoms across time.

| Substance | Predictor | CCEI Depression |  | CCEI Anxiety |  |
| --- | --- | --- | --- | --- | --- |
| | | $\beta$ [95% CI] | p-value | $\beta$ [95% CI] | p-value |
| Alcohol | Main effects |  |  |  |  |
|  | Time | -0.010 [-0.01, -0.009] | <0.001 | -0.011 [-0.011, -0.010] | <0.001 |
|  | Persistent | 0.120 [-0.054, 0.294] | 0.175 | 0.064 [-0.130, 0.258] | 0.518 |
|  | Intermittent | 0.128 [-0.034, 0.290] | 0.122 | 0.117 [-0.065, 0.298] | 0.208 |
|  | Quitters | 0.183 [-0.109, 0.476] | 0.219 | 0.099 [-0.228, 0.426] | 0.551 |
|  | Interaction |  |  |  |  |
|  | Persistent x Time | 0.003 [0.001, 0.004] | <0.001 | 0.003 [0.001, 0.004] | 0.001 |
|  | Intermittent x Time | 0.000 [-0.002, 0.001] | 0.910 | 0.000 [-0.002, 0.001] | 0.690 |
|  | Quitters x Time | 0.000 [-0.003, 0.003] | 0.752 | 0.000 [-0.003, 0.003] | 0.937 |
| Smoking | Main effects |  |  |  |  |
|  | Time | -0.008 [-0.009, -0.008] | <0.001 | -0.009 [-0.01, -0.009] | <0.001 |
|  | Persistent | 0.884 [0.681, 1.087] | <0.001 | 1.051 [0.845, 1.257] | <0.001 |
|  | Intermittent | 0.759 [0.418, 1.100] | <0.001 | 0.860 [0.514, 1.205] | <0.001 |
|  | Quitters | 1.365 [0.528, 2.202] | 0.001 | 1.140 [0.295, 1.985] | 0.008 |
|  | Interaction |  |  |  |  |
|  | Persistent x Time | -0.005 [-0.007, -0.003] | <0.001 | -0.005 [-0.007, -0.003] | <0.001 |
|  | Intermittent x Time | -0.002 [-0.005, 0.001] | 0.192 | -0.003 [-0.006, 0.000] | 0.097 |
|  | Quitters x Time | -0.003 [-0.011, 0.005] | 0.427 | 0.000 [-0.007, 0.008] | 0.926 |

**Note.** Fixed effects beta coefficients ( $\beta$ ) and 95% confidence intervals (CI) are presented for associations between prenatal smoking use patterns (intermittent, persistent, and quitters; reference = abstainers) and trajectories of maternal depressive symptoms measured by the Crown-Crisp Experiential Index (CCEI) and Edinburgh Postnatal Depression Scale (EPDS). Outcomes were assessed during the second and third trimesters of pregnancy and at 8 weeks, 8 months, and 21 months postpartum. All models were adjusted for maternal age, maternal education, marital status, parity, race, and infant sex.

### Smoking Use Patterns and Psychopathology

Compared to women who abstained from smoking during pregnancy, persistent smoking was consistently associated with higher CCEI depressive (β range=0.34–0.75; all p≤0.003) and anxiety symptoms (β range=0.66–0.91; all p<0.001) across pregnancy and the postpartum period (**Table 2**; **Figure 3**), with effect sizes generally attenuating over time. Intermittent smoking was also associated with higher depressive (β range=0.55–0.70; all p≤0.002) and anxiety symptoms (β range=0.44–0.86; all p<0.05) across most assessment periods, though effect sizes were smaller than those observed for persistent smoking. Finally, participants who quit smoking during pregnancy demonstrated elevated depressive symptoms across pregnancy and the early postpartum period (β range=0.85–1.59; p<0.05). Quitters also demonstrated elevated anxiety symptoms in trimester 3 and after 8 months postpartum (β range=1.07–1.30; p<0.05).

**Figure 3.**
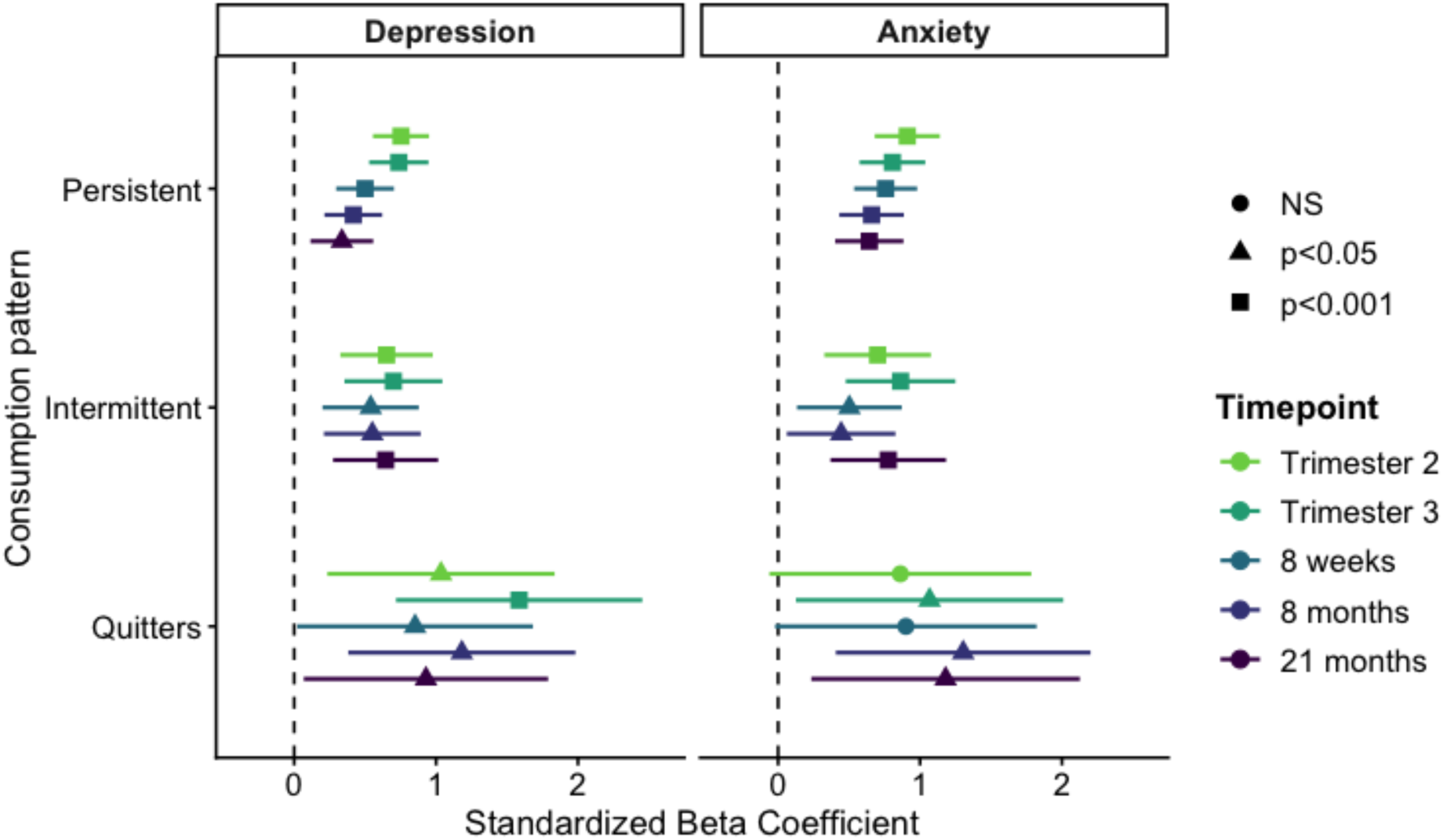
Associations Between Prenatal Smoking Use Patterns and Maternal Depressive and Anxiety Symptoms Across Pregnancy and Postpartum. Points represent standardized regression coefficients (β), and error bars represent 95% confidence intervals. Abstainers/non-smokers served as the reference group. Positive coefficients indicate higher symptom levels relative to abstainers. Depressive and anxiety symptoms were measured with the Crown–Crisp Experiential Index (CCEI). The 8 weeks, 8 months, and 21 months timepoints were measured after birth.

Longitudinal mixed-effects models also identified significant main effects of time on depressive and anxiety symptoms (β range=-0.008 to -0.009; all p<0.001). In addition, persistent, intermittent, and quitting smoking patterns were each associated with higher depressive and anxiety symptom levels than abstinence (**Table 3**). Persistent smoking also significantly interacted with time for both depressive (β=-0.005; p<0.001) and anxiety symptoms (β=-0.005; p<0.001). These findings indicate that although persistent smokers reported higher symptom levels overall, their symptoms declined more rapidly over time than abstainers. No significant interactions with time were observed for intermittent smokers or quitters.

### Sensitivity Analyses

To assess the robustness of findings across measurement approaches, we conducted sensitivity analyses using Edinburgh Postnatal Depression Scale (EPDS) scores as an alternative indicator of depressive symptoms (details in **Supplemental Materials**). Across both alcohol consumption and smoking, the direction, magnitude, and significance of associations were largely consistent with those observed for CCEI depressive symptoms (**Tables S1-S2**). Similarly, correcting for other substance use patterns (i.e., smoking patterns in the alcohol models, and vice versa) yielded nearly identical results to our primary analyses, with no major differences in the direction, magnitude, or significance of associations (**Tables S3-S4**).

## DISCUSSION

This study demonstrates that prenatal substance use is not simply a risk exposure but a meaningful behavioral signal that can help differentiate maternal mental health trajectories across the perinatal period. By modeling alcohol and cigarette use as longitudinal patterns rather than binary indicators, we found that persistence, intermittency, and discontinuation reflect distinct psychological profiles that unfold from pregnancy into the postpartum period. Our results suggest pregnancy represents a key period during which behavioral patterns may signal future vulnerability or adaptation. As such, distinguishing between substance use patterns provides insight into maternal mental health risk that may be obscured by dichotomous exposure models.

Associations between alcohol use patterns and maternal psychopathology were moderate and varied, though still meaningful when examined longitudinally. Persistent alcohol use was associated with elevated depressive and anxiety symptoms beginning in late pregnancy and extending into the postpartum period, whereas intermittent use showed weaker and less stable associations. These findings are consistent with prior literature reporting mixed results for gestational alcohol use, particularly when exposure is treated as binary (Henderson et al., 2007). Importantly, our results suggest that alcohol-related risk may be heightened among women who continue use across pregnancy, rather than among those who reduce or discontinue use (Skagerstróm et al., 2011). Modeling alcohol use patterns over time therefore could help clarify inconsistencies in earlier research and support more precise identification of women who may benefit from targeted screening and early intervention during the perinatal period. In particular, these findings highlight the importance of distinguishing between women who continue substance use across pregnancy and those who reduce or discontinue use, as these groups appear to reflect fundamentally different risk profiles.

By contrast, persistent smoking emerged as the most robust indicator of sustained maternal psychopathology across pregnancy and postpartum. Mothers who continued smoking throughout pregnancy reported higher depressive and anxiety symptoms that persisted well beyond delivery, indicating that smoking persistence may mark ongoing vulnerability rather than transient distress. Importantly, our longitudinal analyses showed that elevated depressive and anxiety symptoms were not limited to women who persistently smoked; women who discontinued or intermittently smoked also demonstrated elevated symptoms relative to abstainers. Although differences between persistent smokers and abstainers narrowed over time, there was no evidence of differential change over time among intermittent smokers or women who discontinued smoking. These findings suggest that changes in smoking behavior do not necessarily coincide with changes in psychological distress, and that smoking patterns during pregnancy may reflect differences in psychological vulnerability that extend beyond smoking status alone. This interpretation is consistent with prior work showing strong associations between prenatal smoking and maternal mental health, as well as evidence that smoking during pregnancy co-occurs with psychiatric comorbidity and socioeconomic disadvantage (Goodwin et al., 2007; Massey et al., 2021). Together, these results suggest persistent, intermittent, and discontinued smoking during pregnancy may function as salient, yet distinct, markers of sustained maternal risk for psychopathology.

A key contribution of this study is the demonstration that discontinuation does not carry the same psychological signature across substances. Women who discontinued alcohol use generally showed depressive and anxiety symptom profiles similar to abstainers, whereas women who discontinued smoking continued to exhibit elevated symptoms across much of the perinatal period. This finding should not necessarily be interpreted to mean that psychological distress impedes smoking cessation. Prior studies comparing pregnancy smokers with pregnancy quitters have found that depression and anxiety do not consistently differentiate these groups after accounting for smoking-related characteristics, and biomarker-validated work has identified pregnancy-specific motivational characteristics associated with cessation even in the absence of differences in depression or anxiety (Massey & Compton, 2012; Massey et al., 2015). Rather, the present findings suggest that behavioral change and psychological vulnerability may be partially dissociable during pregnancy. In other words, women may successfully discontinue smoking while retaining characteristics associated with elevated risk for depression and anxiety. This distinction may help explain why smoking cessation and alcohol cessation identify different maternal mental health profiles and cautions against treating “cessation” as a uniform behavioral phenotype across substances. This finding is particularly important because binary exposure approaches collapse quitters and persistent users into the same categories, potentially obscuring meaningful differences in behavioral and psychological contexts associated with continued versus discontinued use. Recognizing this heterogeneity may improve characterization of maternal mental health risk and inform efforts to identify women who may benefit from additional support during pregnancy and postpartum. The contrasting findings between persistence and discontinuation further suggest that continued substance use during pregnancy may index more entrenched vulnerability, whereas discontinuation may reflect different underlying risk trajectories depending on the substance involved (Forray, 2016). Furthermore, recognizing this heterogeneity is essential for accurately characterizing maternal risk, particularly for clinical and public health efforts aimed at identifying women at risk for sustained depressive and anxiety symptoms during late pregnancy and the early postpartum period.

The observed divergence between alcohol and tobacco also extends earlier evidence that pregnancy does not influence all substance use uniformly. In a prior multisubstance pregnancy cohort, discontinuation varied markedly by substance, with substantially lower cessation of tobacco than alcohol or illicit drugs, while greater depression and anxiety were associated with discontinuation of fewer substance classes among polysubstance users (Massey et al., 2011). The present findings extend that work in a large population-based cohort by demonstrating that substance-specific behavioral patterns are associated with distinct longitudinal trajectories of maternal psychological symptoms.

Our findings also have implications for how pregnancy is conceptualized in studies of substance use and maternal mental health. Pregnancy is not simply a window during which substance exposure occurs; it is also a period of substantial within-person behavioral change. Prior work has shown that smoking can decline even before pregnancy recognition and that pregnancy-associated changes in smoking vary according to pregnancy-specific and parenting-related characteristics (Massey et al., 2022; Level et al., 2023). Viewed in this context, persistence, intermittency, and discontinuation may capture both pre-existing vulnerability and differential behavioral responses to pregnancy. Their associations with subsequent mental health therefore should not be interpreted as evidence that substance-use patterns themselves cause later psychopathology. Rather, these readily observable behavioral trajectories may provide additional information about maternal psychological risk that is lost when substance use is represented at a single time point.

These findings may also have practical implications for perinatal mental health assessment. Substance-use screening is already clinically relevant during pregnancy, but the present findings suggest that a single assessment of current use may provide less information than asking how use has changed since conception. Persistent or intermittent smoking, and persistent alcohol use, identified women with elevated depressive and anxiety symptoms extending into the postpartum period. Importantly, smoking cessation did not identify the same low-symptom profile as alcohol cessation, suggesting that successful discontinuation of smoking should not be interpreted as evidence that co-occurring psychological vulnerability has resolved. Thus, brief characterization of whether substance use has persisted, fluctuated, or stopped during pregnancy could complement—not replace—direct screening for depression and anxiety and may identify women for whom continued mental health assessment across pregnancy and postpartum is particularly warranted.

Some limitations should be considered when interpreting these findings. First, substance use and mental health symptoms were assessed via self-report, which may be subject to underreporting or recall bias. Future studies could strengthen measurement by incorporating objective indicators of substance use, such as biological assays, alongside self-report measures, as well as using more frequent or real-time assessments to better capture mental health symptoms across the perinatal period. Second, the observational design precludes causal inference, and residual confounding cannot be ruled out despite adjustment for key sociodemographic factors.

In addition, depressive and anxiety symptoms were already present during pregnancy while substance-use trajectories were unfolding, precluding clear temporal ordering. The observed associations may therefore reflect effects of psychological symptoms on substance use, effects of substance use on symptoms, shared antecedent vulnerability, pregnancy-associated processes influencing both, or some combination of these pathways. Third, the ALSPAC cohort reflects pregnancies in the United Kingdom during the early 1990s, representing a substantial temporal gap from contemporary perinatal contexts in which public health awareness of prenatal substance use and maternal mental health has increased, particularly around alcohol use during pregnancy. Finally, ALSPAC is predominantly White, limiting generalizability to more diverse populations. Although the longitudinal design, repeated assessments across pregnancy, and extended postpartum follow-up strengthen confidence in the observed patterns, replication in contemporary and more diverse cohorts is needed.

## CONCLUSIONS

This study shows that longitudinal patterns of prenatal alcohol use and smoking are differentially associated with maternal depressive and anxiety symptoms across pregnancy and the postpartum period. By moving beyond binary exposure definitions, we demonstrate that persistence, intermittency, and discontinuation identify meaningfully different maternal mental health profiles—and that these patterns differ by substance. Persistent use was associated with sustained psychological distress, whereas discontinuation was associated with different psychological profiles for alcohol and smoking. These findings highlight the value of characterizing maternal substance-use behavior longitudinally rather than treating prenatal exposure as a static binary condition and may improve identification of women who would benefit from additional mental health support across the transition to parenthood. Ultimately, incorporating pattern-based approaches into perinatal research may improve precision in identifying women at elevated risk and strengthen efforts to support maternal mental health across the transition to parenthood.

## ACKNOWLEDGEMENTS

This work was supported by the National Institute on Alcohol Abuse and Alcoholism (NIAAA; R21AA030640 awarded to AAL) and the National Institute on Drug Abuse (NIDA; R01DA050700 awarded to SHM) of the National Institutes of Health. The content is solely the responsibility of the authors and does not necessarily represent the official views of the National Institutes of Health. AAL is supported by an MQ Fellows Award from the MQ Foundation (MQF22\9).

We are extremely grateful to all the families who took part in this study, the midwives for their help in recruiting them, and the whole ALSPAC team, which includes data collection staff, data and administration staff, technical managers and the technical staff with the Bristol Bioresource Laboratory, based within the University of Bristol. The UK Medical Research Council and Welcome (Grant ref: MR/Z505924/1) and the University of Bristol provide core support for ALSPAC. This publication is the work of the authors, who will serve as guarantors for the contents of this paper.

## AUTHOR CONTRIBUTIONS

KNP helped conceptualize the study ran the linear regression analyses, interpreted the findings, and wrote the manuscript. KYTL assisted in the design and interpretation of the study and provided critical input in writing the manuscript. SHM helped conceptualize the study and provided critical input on the results and manuscript. AAL obtained grant support for this work, conceptualized the study, ran the linear mixed-effects analyses, interpreted the results, and helped write the manuscript.

## DATA AVAILABILITY

The informed consent obtained from ALSPAC participants does not allow the data to be made available through any third-party maintained public repository. All data are available by request from the ALSPAC Executive Committee for researchers who meet the criteria for access to confidential data (https://bristol.ac.uk/alspac/researchers/access). The ALSPAC study website contains details of all available data (https://www.bristol.ac.uk/alspac/researchers/our-data/).

## INCLUSION AND ETHICS

Ethical approval for the study was obtained from the ALSPAC Ethics and Law Committee and the Local Research Ethics Committees. Informed consent for the use of data collected via questionnaires and clinics was obtained from participants following the recommendations of the ALSPAC Ethics and Law Committee. Study participation was voluntary, and participants provided informed consent for the intended use of all data collected during collection sweeps. The completion of a questionnaire, either on paper or online, was considered to be written consent from participants to use their data for research purposes. Data in this study were fully anonymized, and participants can contact the study team at any time to retrospectively withdraw consent for their data to be used. Secondary analyses of these data were approved with oversight by the Mass General Brigham Institutional Review Boards (Protocol 2023P000011).

