## Supplemental materials for "Patterns of substance use during pregnancy are differentially associated with maternal mental health across the perinatal period"

**SUPPLEMENTAL METHODS**

*Recoding of alcohol use variables*

Prospective alcohol consumption was reported by mothers via questionnaires at study intake and repeated in each subsequent trimester of pregnancy until birth. Participants were advised that one drink was equivalent to 10 grams of pure alcohol, a half-pint of beer/lager, one glass of wine, and one pub-measure of spirits or other alcoholic drinks. Responses were collected in ALSPAC waves A, B, and C, each wave containing reports from a mixture of gestational stages after conception (Figure S1). As such, we used week of gestation at questionnaire completion to assign prospective alcohol consumption reports to pregnancy trimesters (<18 weeks: first trimester; 18-27 weeks: second trimester; $>$27 weeks: third trimester). All postpartum responses are treated as retrospective reports and excluded from analyses.

*Recoding of smoking variables*

Prospective smoking status and/or quantities during pregnancy were reported by mothers during ALSPAC waves A, B, C, each containing reports from a mixture of gestational stages after conception in the same manner as maternally-reported alcohol consumption levels. Therefore, we applied the same timepoint-recoding procedures as with PAE reports, to create dichotomous maternal smoking variables for each trimester (0/1). To maximize the availability of responses for the third trimester, we used retrospective smoking reports obtained up to 12 weeks postpartum, and the rest were excluded from analyses.

*Covariates*

All covariates were selected a priori based on existing literature and were included to reduce potential confounding in the associations between prenatal substance use patterns and maternal psychopathology. Prior research indicates that maternal sociodemographic characteristics are strongly associated with patterns of alcohol and cigarette use during pregnancy, as well as with risk for depressive and anxiety symptoms across the perinatal period (Forray, 2016; Liu et al., 2021). Parity and marital status have similarly been linked to variability in both prenatal substance use and maternal mental health outcomes, reflecting broader relational and caregiving contexts (Qiu et al., 2022). Infant sex was included given mixed evidence regarding sex-specific associations with maternal psychological symptoms in the postpartum period. We coded the covariates in the following way:

- *maternal age at delivery* – categorical variable with three categories of response, ages 15-19, ages 20-35, and age 36+
- *maternal education* – categorical variable with four categories of response, less than O-level, O-level, A-level, and degree or above.
- *marital status* – categorical variable with three categories of response, married, never married, widowed/divorced/separated.
- *number of previous pregnancies (parity)* – ordinal variable of 0, 1, 2, 3+ previous children.
- *race/ethnicity* – binary variable of non-white/white (0/1) due to the low frequency of non-white participants in ALSPAC. Race/ethnicity was determined based on maternal self-report at birth.
- *infant sex* – binary variable of male/female (1/2)

*Edinburgh Postnatal Depression Scale*

Mother also reported depressive symptoms at the same five timepoints using the Edinburgh Postnatal Depression Scale (EPDS). Briefly, the EPDS is a 10-item questionnaire designed to assess the frequency of depressive symptoms experienced over the past week, including low mood, anhedonia, guilt, and sleep disturbance. Items are rated on a four-point Likert scale ranging from 0 to 3, yielding total scores from 0 to 30, with higher scores indicating greater depressive symptom severity. We used the EPDS in sensitivity analyses to determine whether the measurement scale impacted our results.

**SUPPLEMENTAL TABLES**

**Table S1: Sensitivity analysis of associations between prenatal substance use patterns and maternal depressive and anxiety symptoms across pregnancy and the postpartum period using the CCEI and EPDS**

| **Substance** | **Consumption pattern** | **Outcome timepoint** | **CCEI depression** | | | **EPDS depression** | | |
| --- | --- | --- | --- | --- | --- | --- | --- | --- |
|  |  |  | **N** | **β [95% CI]** | **p-value** | **N** | **β [95% CI]** | **p-value** |
| Alcohol | Persistent | Trimester 2 | 7,629 | 0.14 [-0.04, 0.33] | 0.125 | 7,716 | 0.29 [0.01, 0.57] | 0.045 |
|  |  | Trimester 3 | 7,661 | 0.23 [0.03, 0.42] | 0.023 | 7,823 | 0.46 [0.16, 0.76] | 0.002 |
|  |  | 8 weeks | 7,351 | 0.34 [0.15, 0.53] | <0.001 | 7,435 | 0.64 [0.35, 0.93] | <0.001 |
|  |  | 8 months | 7,169 | 0.37 [0.18, 0.55] | <0.001 | 7,214 | 0.43 [0.14, 0.72] | 0.004 |
|  |  | 21 months | 6,644 | 0.40 [0.20, 0.60] | <0.001 | 6,684 | 0.53 [0.23, 0.84] | <0.001 |
|  | Intermittent | Trimester 2 | 7,629 | 0.11 [-0.06, 0.28] | 0.203 | 7,716 | 0.21 [-0.05, 0.48] | 0.115 |
|  |  | Trimester 3 | 7,661 | 0.12 [-0.06, 0.30] | 0.186 | 7,823 | 0.15 [-0.13, 0.43] | 0.294 |
|  |  | 8 weeks | 7,351 | 0.21 [0.03, 0.38] | 0.019 | 7,435 | 0.24 [-0.03, 0.52] | 0.077 |
|  |  | 8 months | 7,169 | 0.20 [0.03, 0.37] | 0.024 | 7,214 | 0.18 [-0.09, 0.45] | 0.197 |
|  |  | 21 months | 6,644 | 0.11 [-0.08, 0.29] | 0.262 | 6,684 | 0.05 [-0.24, 0.34] | 0.745 |
|  | Quitters | Trimester 2 | 7,629 | 0.07 [-0.23, 0.38] | 0.636 | 7,716 | 0.26 [-0.21, 0.74] | 0.279 |
|  |  | Trimester 3 | 7,661 | 0.23 [-0.1, 0.56] | 0.171 | 7,823 | 0.24 [-0.26, 0.74] | 0.342 |
|  |  | 8 weeks | 7,351 | 0.22 [-0.1, 0.54] | 0.173 | 7,435 | 0.45 [-0.04, 0.94] | 0.073 |
|  |  | 8 months | 7,169 | 0.37 [0.06, 0.68] | 0.020 | 7,214 | 0.62 [0.13, 1.11] | 0.013 |
|  |  | 21 months | 6,644 | 0.09 [-0.24, 0.43] | 0.578 | 6,684 | 0.18 [-0.34, 0.70] | 0.495 |
| Smoking | Persistent | Trimester 2 | 7,629 | 0.75 [0.55, 0.95] | <0.001 | 7,716 | 1.52 [1.22, 1.82] | <0.001 |
|  |  | Trimester 3 | 7,661 | 0.74 [0.53, 0.95] | <0.001 | 7,823 | 1.26 [0.94, 1.58] | <0.001 |
|  |  | 8 weeks | 7,351 | 0.50 [0.30, 0.70] | <0.001 | 7,435 | 1.12 [0.8, 1.44] | <0.001 |
|  |  | 8 months | 7,169 | 0.42 [0.21, 0.62] | <0.001 | 7,214 | 0.91 [0.59, 1.22] | <0.001 |
|  |  | 21 months | 6,644 | 0.34 [0.12, 0.56] | 0.003 | 6,684 | 0.93 [0.58, 1.27] | <0.001 |
|  | Intermittent | Trimester 2 | 7,629 | 0.65 [0.33, 0.98] | <0.001 | 7,716 | 1.16 [0.66, 1.66] | <0.001 |
|  |  | Trimester 3 | 7,661 | 0.70 [0.35, 1.04] | <0.001 | 7,823 | 1.02 [0.49, 1.54] | <0.001 |
|  |  | 8 weeks | 7,351 | 0.54 [0.20, 0.88] | 0.002 | 7,435 | 0.69 [0.16, 1.22] | 0.011 |
|  |  | 8 months | 7,169 | 0.55 [0.21, 0.89] | 0.002 | 7,214 | 0.67 [0.14, 1.21] | 0.013 |
|  |  | 21 months | 6,644 | 0.64 [0.27, 1.01] | <0.001 | 6,684 | 1.14 [0.56, 1.71] | <0.001 |
|  | Quitters | Trimester 2 | 7,629 | 1.03 [0.23, 1.84] | 0.011 | 7,716 | 2.42 [1.2, 3.65] | <0.001 |
|  |  | Trimester 3 | 7,661 | 1.59 [0.72, 2.45] | <0.001 | 7,823 | 2.46 [1.13, 3.78] | <0.001 |
|  |  | 8 weeks | 7,351 | 0.85 [0.02, 1.68] | 0.044 | 7,435 | 1.84 [0.53, 3.16] | 0.006 |
|  |  | 8 months | 7,169 | 1.18 [0.38, 1.98] | 0.004 | 7,214 | 1.77 [0.52, 3.03] | 0.006 |
|  |  | 21 months | 6,644 | 0.93 [0.07, 1.79] | 0.035 | 6,684 | 2.77 [1.43, 4.1] | <0.001 |

**Note.** Standardized beta coefficients (β) and 95% confidence intervals (CI) are presented for associations between prenatal smoking use patterns (intermittent, persistent, and quitters; reference = abstainers) and maternal depressive symptoms measured by the Crown-Crisp Experiential Index (CCEI) and Edinburgh Postnatal Depression Scale (EPDS). Outcomes were assessed during the second and third trimesters of pregnancy and at 8 weeks, 8 months, and 21 months postpartum. All models were adjusted for maternal age, maternal education, marital status, parity, race, and infant sex.

**Table S2: Sensitivity analysis of associations between prenatal substance use patterns and trajectories of maternal depressive and anxiety symptoms using the CCEI and EPDS**

| **Substance** | **Predictor** | **CCEI Depression** | | **EPDS Depression** | |
| --- | --- | --- | --- | --- | --- |
|  |  | **β [95% CI]** | **p-value** | **β [95% CI]** | **p-value** |
| Alcohol | Main effects |  |  |  |  |
|  | Time | -0.010 [-0.01, -0.009] | <0.001 | -0.011 [-0.012, -0.01] | <0.001 |
|  | Persistent | 0.120 [-0.054, 0.294] | 0.175 | 0.324 [0.056, 0.592] | 0.018 |
|  | Intermittent | 0.128 [-0.034, 0.290] | 0.122 | 0.190 [-0.060, 0.440] | 0.136 |
|  | Quitters | 0.183 [-0.109, 0.476] | 0.219 | 0.291 [-0.159, 0.74] | 0.205 |
|  | Interaction |  |  |  |  |
|  | Persistent x Time | 0.003 [0.001, 0.004] | <0.001 | 0.002 [0.000, 0.004] | 0.100 |
|  | Intermittent x Time | 0.000 [-0.002, 0.001] | 0.910 | -0.001 [-0.003, 0.001] | 0.364 |
|  | Quitters x Time | 0.000 [-0.003, 0.003] | 0.752 | 0.001 [-0.003, 0.005] | 0.775 |
| Smoking | Main effects |  |  |  |  |
|  | Time | -0.008 [-0.009, -0.008] | <0.001 | -0.010 [-0.011, -0.009] | <0.001 |
|  | Persistent | 0.884 [0.681, 1.087] | <0.001 | 1.612 [1.328, 1.896] | <0.001 |
|  | Intermittent | 0.759 [0.418, 1.100] | <0.001 | 1.174 [0.698, 1.650] | <0.001 |
|  | Quitters | 1.365 [0.528, 2.202] | 0.001 | 2.402 [1.237, 3.567] | <0.001 |
|  | Interaction |  |  |  |  |
|  | Persistent x Time | -0.005 [-0.007, -0.003] | <0.001 | -0.008 [-0.01, -0.005] | <0.001 |
|  | Intermittent x Time | -0.002 [-0.005, 0.001] | 0.192 | -0.004 [-0.008, 0.001] | 0.114 |
|  | Quitters x Time | -0.003 [-0.011, 0.005] | 0.427 | -0.001 [-0.012, 0.009] | 0.799 |

**Note.** Fixed effects beta coefficients (β) and 95% confidence intervals (CI) are presented for associations between prenatal smoking use patterns (intermittent, persistent, and quitters; reference = abstainers) and trajectories of maternal depressive symptoms measured by the Crown-Crisp Experiential Index (CCEI) and Edinburgh Postnatal Depression Scale (EPDS). Outcomes were assessed during the second and third trimesters of pregnancy and at 8 weeks, 8 months, and 21 months postpartum. All models were adjusted for maternal age, maternal education, marital status, parity, race, and infant sex.

**Table S3.** **Sensitivity analysis of associations between prenatal substance use patterns and maternal depressive and anxiety symptoms when correcting for other substance use patterns.**

| **Substance** | **Consumption pattern** | **Outcome timepoint** | **CCEI Depression** | | | **CCEI Anxiety** | | |
| --- | --- | --- | --- | --- | --- | --- | --- | --- |
|  |  |  | **N** | **β [95% CI]** | **p-value** | **N** | **β [95% CI]** | **p-value** |
| Alcohol | Persistent | Trimester 2 | 7,629 | 0.12 [-0.07, 0.30] | 0.214 | 7,663 | 0.09 [-0.13, 0.30] | 0.424 |
|  |  | Trimester 3 | 7,661 | 0.21 [0.01, 0.40] | 0.039 | 7,675 | 0.13 [-0.08, 0.35] | 0.220 |
|  |  | 8 weeks | 7,351 | 0.32 [0.14, 0.51] | <0.001 | 7,364 | 0.29 [0.09, 0.49] | 0.005 |
|  |  | 8 months | 7,169 | 0.35 [0.17, 0.54] | <0.001 | 7,195 | 0.25 [0.04, 0.46] | 0.017 |
|  |  | 21 months | 6,644 | 0.38 [0.19, 0.58] | <0.001 | 6,597 | 0.30 [0.09, 0.52] | 0.006 |
|  | Intermittent | Trimester 2 | 7,629 | 0.1 [-0.07, 0.28] | 0.230 | 7,663 | 0.10 [-0.09, 0.30] | 0.306 |
|  |  | Trimester 3 | 7,661 | 0.11 [-0.07, 0.29] | 0.216 | 7,675 | 0.10 [-0.10, 0.3] | 0.347 |
|  |  | 8 weeks | 7,351 | 0.20 [0.03, 0.37] | 0.023 | 7,364 | 0.20 [0.01, 0.39] | 0.041 |
|  |  | 8 months | 7,169 | 0.19 [0.02, 0.37] | 0.028 | 7,195 | 0.03 [-0.16, 0.22] | 0.770 |
|  |  | 21 months | 6,644 | 0.10 [-0.09, 0.28] | 0.300 | 6,597 | 0.04 [-0.16, 0.24] | 0.717 |
|  | Quitters | Trimester 2 | 7,629 | 0.05 [-0.25, 0.36] | 0.739 | 7,663 | 0.08 [-0.28, 0.43] | 0.674 |
|  |  | Trimester 3 | 7,661 | 0.21 [-0.12, 0.54] | 0.208 | 7,675 | 0.03 [-0.33, 0.39] | 0.882 |
|  |  | 8 weeks | 7,351 | 0.21 [-0.11, 0.52] | 0.205 | 7,364 | 0.04 [-0.31, 0.38] | 0.839 |
|  |  | 8 months | 7,169 | 0.36 [0.04, 0.67] | 0.025 | 7,195 | 0.19 [-0.16, 0.54] | 0.289 |
|  |  | 21 months | 6,644 | 0.08 [-0.25, 0.42] | 0.632 | 6,597 | -0.03 [-0.40, 0.33] | 0.856 |
| Smoking | Persistent | Trimester 2 | 7,629 | 0.75 [0.55, 0.95] | <0.001 | 7,663 | 0.91 [0.68, 1.14] | <0.001 |
|  |  | Trimester 3 | 7,661 | 0.73 [0.52, 0.94] | <0.001 | 7,675 | 0.80 [0.57, 1.03] | <0.001 |
|  |  | 8 weeks | 7,351 | 0.49 [0.28, 0.69] | <0.001 | 7,364 | 0.75 [0.53, 0.97] | <0.001 |
|  |  | 8 months | 7,169 | 0.40 [0.20, 0.61] | <0.001 | 7,195 | 0.65 [0.42, 0.88] | <0.001 |
|  |  | 21 months | 6,644 | 0.32 [0.1, 0.54] | 0.004 | 6,597 | 0.63 [0.39, 0.87] | <0.001 |
|  | Intermittent | Trimester 2 | 7,629 | 0.65 [0.32, 0.97] | <0.001 | 7,663 | 0.70 [0.32, 1.07] | <0.001 |
|  |  | Trimester 3 | 7,661 | 0.69 [0.34, 1.03] | <0.001 | 7,675 | 0.86 [0.47, 1.24] | <0.001 |
|  |  | 8 weeks | 7,351 | 0.53 [0.19, 0.87] | 0.002 | 7,364 | 0.50 [0.13, 0.86] | 0.009 |
|  |  | 8 months | 7,169 | 0.54 [0.20, 0.88] | 0.002 | 7,195 | 0.44 [0.05, 0.82] | 0.026 |
|  |  | 21 months | 6,644 | 0.63 [0.26, 1.01] | <0.001 | 6,597 | 0.77 [0.36, 1.18] | <0.001 |
|  | Quitters | Trimester 2 | 7,629 | 1.03 [0.23, 1.83] | 0.011 | 7,663 | 0.86 [-0.06, 1.78] | 0.068 |
|  |  | Trimester 3 | 7,661 | 1.59 [0.72, 2.45] | <0.001 | 7,675 | 1.06 [0.12, 2.01] | 0.027 |
|  |  | 8 weeks | 7,351 | 0.84 [0.01, 1.67] | 0.048 | 7,364 | 0.88 [-0.04, 1.81] | 0.060 |
|  |  | 8 months | 7,169 | 1.18 [0.38, 1.98] | 0.004 | 7,195 | 1.31 [0.41, 2.20] | 0.004 |
|  |  | 21 months | 6,644 | 0.92 [0.06, 1.78] | 0.037 | 6,597 | 1.17 [0.22, 2.11] | 0.015 |

**Note.** Standardized beta coefficients (β) and 95% confidence intervals (CI) are presented for associations between prenatal smoking use patterns (intermittent, persistent, and quitters; reference = abstainers) and maternal depressive symptoms measured by the Crown-Crisp Experiential Index (CCEI) and Edinburgh Postnatal Depression Scale (EPDS). Outcomes were assessed during the second and third trimesters of pregnancy and at 8 weeks, 8 months, and 21 months postpartum. All models were adjusted for maternal age, maternal education, marital status, parity, race, and infant sex. In addition, we corrected for smoking patterns in the alcohol models and alcohol patterns in the smoking models.

**Table S4. Sensitivity analysis of associations between prenatal substance use patterns and maternal depressive symptoms across pregnancy when correcting for other substance use patterns.**

| **Substance** | **Predictor** | **CCEI Depression** | | **CCEI Anxiety** | |
| --- | --- | --- | --- | --- | --- |
|  |  | **β [95% CI]** | **p-value** | **β [95% CI]** | **p-value** |
| Alcohol | Main effects |  |  |  |  |
|  | Time | -0.010 [-0.010, -0.009] | <0.001 | -0.011 [-0.011, -0.010] | <0.001 |
|  | Persistent | 0.100 [-0.074, 0.273] | 0.260 | 0.037 [-0.157, 0.230] | 0.711 |
|  | Intermittent | 0.122 [-0.040, 0.283] | 0.140 | 0.109 [-0.072, 0.289] | 0.237 |
|  | Quitters | 0.168 [-0.124, 0.459] | 0.259 | 0.078 [-0.248, 0.403] | 0.639 |
|  | Interaction |  |  |  |  |
|  | Persistent x Time | 0.003 [0.001, 0.004] | <0.001 | 0.003 [0.001, 0.004] | 0.001 |
|  | Intermittent x Time | 0.000 [-0.002, 0.001] | 0.901 | 0.000 [-0.002, 0.001] | 0.680 |
|  | Quitters x Time | 0.000 [-0.002, 0.003] | 0.754 | 0.000 [-0.003, 0.003] | 0.938 |
| Smoking | Main effects |  |  |  |  |
|  | Time | -0.008 [-0.009, -0.008] | <0.001 | -0.009 [-0.01, -0.009] | <0.001 |
|  | Persistent | 0.874 [0.671, 1.077] | <0.001 | 1.044 [0.837, 1.250] | <0.001 |
|  | Intermittent | 0.750 [0.409, 1.091] | <0.001 | 0.854 [0.508, 1.200] | <0.001 |
|  | Quitters | 1.369 [0.531, 2.206] | 0.001 | 1.139 [0.294, 1.984] | 0.008 |
|  | Interaction |  |  |  |  |
|  | Persistent x Time | -0.005 [-0.007, -0.003] | <0.001 | -0.005 [-0.007, -0.003] | <0.001 |
|  | Intermittent x Time | -0.002 [-0.005, 0.001] | 0.194 | -0.003 [-0.006, 0.000] | 0.097 |
|  | Quitters x Time | -0.003 [-0.011, 0.004] | 0.423 | 0.000 [-0.007, 0.008] | 0.929 |

**Note.** Fixed effects beta coefficients (β) and 95% confidence intervals (CI) are presented for associations between prenatal smoking use patterns (intermittent, persistent, and quitters; reference = abstainers) and trajectories of maternal depressive symptoms measured by the Crown-Crisp Experiential Index (CCEI). Outcomes were assessed during the second and third trimesters of pregnancy and at 8 weeks, 8 months, and 21 months postpartum. All models were adjusted for maternal age, maternal education, marital status, parity, race, and infant sex. In addition, we corrected for smoking patterns in the alcohol models and alcohol patterns in the smoking models.

**SUPPLEMENTAL FIGURES**

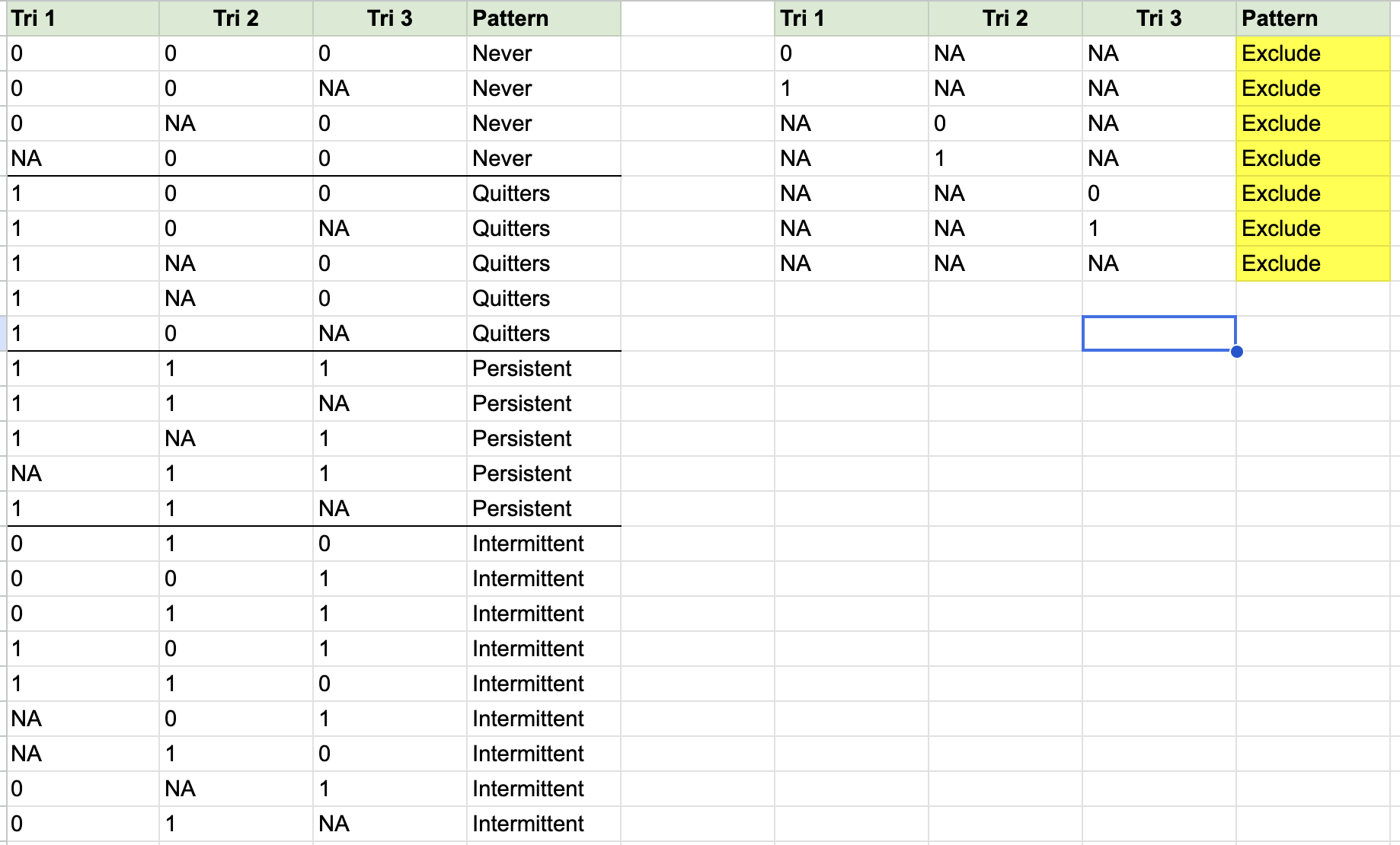

**Figure S1. Coding of substance use patterns.** This coding was implemented to identify the four patterns of prenatal substance use for both alcohol and smoking (separate variables), which include abstainers/never, quitters, persistent users, and intermittent users.
